# Formation of oxidized gases and secondary organic aerosol from a commercial oxidant-generating electronic air cleaner

**DOI:** 10.1101/2021.06.01.21258186

**Authors:** Taekyu Joo, Jean C. Rivera-Rios, Daniel Alvarado-Velez, Sabrina Westgate, Nga Lee Ng

## Abstract

Airborne virus transmission during the COVID-19 pandemic increased the demand for indoor air cleaners. While some commercial electronic air cleaners could be effective in reducing primary pollutants and inactivating bioaerosol, studies on the formation of secondary products from oxidation chemistry during their use are limited. Here, we measured oxygenated volatile organic compounds (OVOCs) and the chemical composition of particles generated from a hydroxyl radical generator in an office. During operation, enhancements in OVOCs, especially low-molecular-weight organic and inorganic acids, were detected. Rapid increases in particle number and volume concentrations were observed, corresponding to the formation of highly-oxidized secondary organic aerosol (SOA) (O:C ∼1.3). The organic mass spectra showed an enhanced signal at *m/z* 44 (CO^2+^) and the aerosol evolved with a slope of ∼ -1 in the Van Krevelen diagram. These results suggest that organic acids generated during VOC oxidation contributed to particle nucleation and SOA formation. Nitrate, sulfate, and chloride also increased during the oxidation without a corresponding increase in ammonium, suggesting organic nitrate, organic sulfate, and organic chloride formation. As secondary species are reported to have detrimental health effects, further studies are needed to evaluate potential OVOCs and SOA formation from electronic air cleaners in different indoor environments.

**Synopsis:** We observed formation of oxygenated volatile organic compounds and secondary organic aerosol from an electronic air cleaner.

## Introduction

People spend most of their time indoors, making the air quality in these spaces an important factor for human health. Indoor air quality (IAQ) depends on several factors, including but not limited to: exchange with outdoor air, filtration, emissions from indoor sources, chemical reactions i.e., via oxidation or multi-phase processes, and deposition onto surfaces.^1^ Due to the impact of IAQ on health, there is a growing demand for air cleaning technologies meant to reduce exposure to potentially detrimental substances indoors. This demand has increased considerably during the course of the recent COVID-19 (severe acute respiratory syndrome coronavirus 2, SARS-CoV-2) pandemic due to the increased recognition of the role of airborne virus transmission, especially indoors.^2-6^

Air cleaners are usually deployed with the intention to remove indoor pollutants such as particles or volatile organic compounds (VOCs), as well as to inactivate pathogens. Two types of air cleaning technologies are commonly used to remove particles: mechanical filtration and electronic air cleaners (e.g., ionizers and electrostatic precipitators). Gaseous pollutants such as odors and VOCs can be removed via a number of different technologies: adsorbent media air filters (e.g., activated carbon) and various electronic air cleaning devices such as photocatalytic oxidation (PCO), plasma, and ozone-generating equipment among others.^7-9^ In addition, hydroxyl radical (OH) generation via photolysis of ozone or water is also used to destroy odors and VOCs, usually as a substitute for ozone-generating air cleaners.^10, 11^ Among these cleaning technologies, ultraviolet germicidal irradiation (UVGI), ionizers, ozone oxidations, and PCO purifiers have been shown to be capable of inactivating viruses, bacteria, and other bioaerosol.^6, 8, 9, 12-19^

There are increasing concerns regarding the use of electronic air cleaners as these devices can potentially generate unintended byproducts via oxidation chemistry similar to that in the atmosphere.^20, 21^ The oxidation mechanism of VOCs in the atmosphere can be simplified as the following: (1) initial attack of the VOCs by oxidants (OH, O_3_, and NO_3_), (2) organic peroxy radical reactions, and in some cases (3) alkoxy radical reactions.^22, 23^ Organic peroxy radicals can react with other species in the atmosphere (e.g., NO, NO_2_, HO_2_, etc.) and undergo functionalization or form alkoxy radicals. Alkoxy radicals can fragment and form smaller organic compounds in the atmosphere that can be oxidized further. Fragmentation leads to increased volatility whereas functionalization decreases volatility and increases solubility.^22^ These complex, multi-generational, gas-phase oxidation processes result in the formation of a large variety of organic compounds, which can undergo gas-particle partitioning and/or nucleation to form secondary organic aerosol (SOA). While some byproducts of VOC oxidation can have adverse health effects,^24-29^ systematic investigations of potential formation of organic gases and aerosol during the operation of oxidant/ion-generating air cleaners indoors are scarce. Previous studies are limited to investigating the formation of ozone, NO_x_, CO, CO_2_, less-oxidized VOCs, or particle number and mass concentrations, but not on the composition of more-oxidized VOCs or aerosol.^8, 9, 30^

In this work, we evaluated the effect of a commercial electronic air cleaner (hydroxyl radical generator) operated inside an office. We monitored gas-phase oxidized products and PM_1_ (particulate matter less than 1 µm in diameter) size distribution and composition. We show that the operation of this device leads to the formation of small organic acids and increases PM_1_ number and mass concentrations. These results show that care must be taken when choosing an adequate and appropriate air cleaning technology for a particular environment and task.

## Materials and methods

The experiment was performed in an office (∼ 16 m^2^) in the Ford Environmental Sciences and Technology Building at the Georgia Institute of Technology. We performed the experiment in the following sequence: 1) 2.33 hours of office background sampling, 2) 1.5 hours of hydroxyl generator operation (Titan Model #4000, International Ozone Technologies Group, Inc., Delray Beach, FL), and 3) 1.5 hours of sampling after the device was turned off. Briefly, the device generates OH radical and hydrogen peroxide (H_2_O_2_) via photocatalytic reaction of TiO_2_ with UV-A range (365 - 385 nm) light and H_2_O and O_2_ in the air.^31, 32^ Many other brands of hydroxyl generator are available in the market and employ a similar technology.

Gas-phase organic compounds were measured and reported as counts per second using a high-resolution time-of-flight chemical ionization mass spectrometer (HR-ToF-CIMS, Aerodyne Research Inc., Billerica, MA) with iodide (I^-^) as a reagent ion, which selectively measures oxygenated organics.^33^ O_3_ and NO_x_ were monitored using an O_3_ Analyzer (T400, Teledyne, City of Industry, CA), a NO-NO_2_-NO_x_ Analyzer (42C, Thermo Fisher Scientific, Waltham, MA), and a Cavity Attenuated Phase Shift NO_2_ monitor (CAPS, Aerodyne Inc.).

Size-resolved PM_1_ number and volume concentrations were measured using a scanning mobility particle sizer (SMPS). The SMPS is a combination of a differential mobility analyzer (DMA) (TSI 3040, TSI Inc., Shoreview, MN) and a condensation particle counter (CPC) (TSI 3775). In addition, we deployed a separate CPC (TSI 3025 A) to monitor the total number concentration of particles (all particles under roughly 3 µm). Aerosol chemical composition was monitored using a high-resolution time-of-flight aerosol mass spectrometer (HR-ToF-AMS, Aerodyne Research Inc.). HR-ToF-AMS quantifies organics, nitrate, sulfate, ammonium, and chloride mass concentrations and measures the bulk elemental composition of the particles (e.g., O:C and H:C ratios).^34, 35^ The elemental ratios for particles were calculated based on the “Improved-Ambient” method.^35^

## Results

### Formation of oxidized VOCs (OVOCs)

The immediate formation of oxygenated products was observed by the HR-ToF-CIMS (Figure 1) when the device was turned on. Formic acid (*m/z* 173, CH_2_O_2_I^-^), acetic acid (*m/z* 187, C_2_H_4_O_2_I^-^), iminoacetic acid (*m/z* 200, C_2_H_3_NO_2_I^-^), oxamide (*m/z* 215, C_2_H_4_N_2_O_2_I^-^), glyceraldehyde (*m/z* 217, C_3_H_6_O_3_I^-^), glycerol (*m/z* 219, C_3_H_8_O_3_I^-^), alanine (*m/z* 216, C_3_H_7_NO_2_I^-^), and acetoacetic acid (*m/z* 229, C_4_H_6_O_3_I^-^) are identified and showed the most obvious enhancements during the operation period. Enhanced glyceraldehyde and glycerol at the beginning of the experiment (12:10 pm) was likely due to the presence of people in the office initially (to set up instruments for this study), as these compounds are formed as intermediates in metabolism and widely used in cosmetics or as an additive in foods.^36-38^ Nitrous acid (*m/z* 174, HONOI^-^), which is an inorganic acid, also increased during the operation. Hydrogen peroxide (*m/z* 161, H_2_O_2_I^-^) increased during the background period and decreased during the operation of device. As mentioned in the previous section, TiO_2_ photocatalytic technology is reported to produce H_2_O_2_ as another product.^31^ However, H_2_O_2_ decreased when the device was turned on and rebounded after the device was turned off. The pre-existing H_2_O_2_ in the office could have been interacting with the generated OH radical but flattened as a result of regeneration via self-reaction of hydroperoxyl radical.

**Figure 1.**
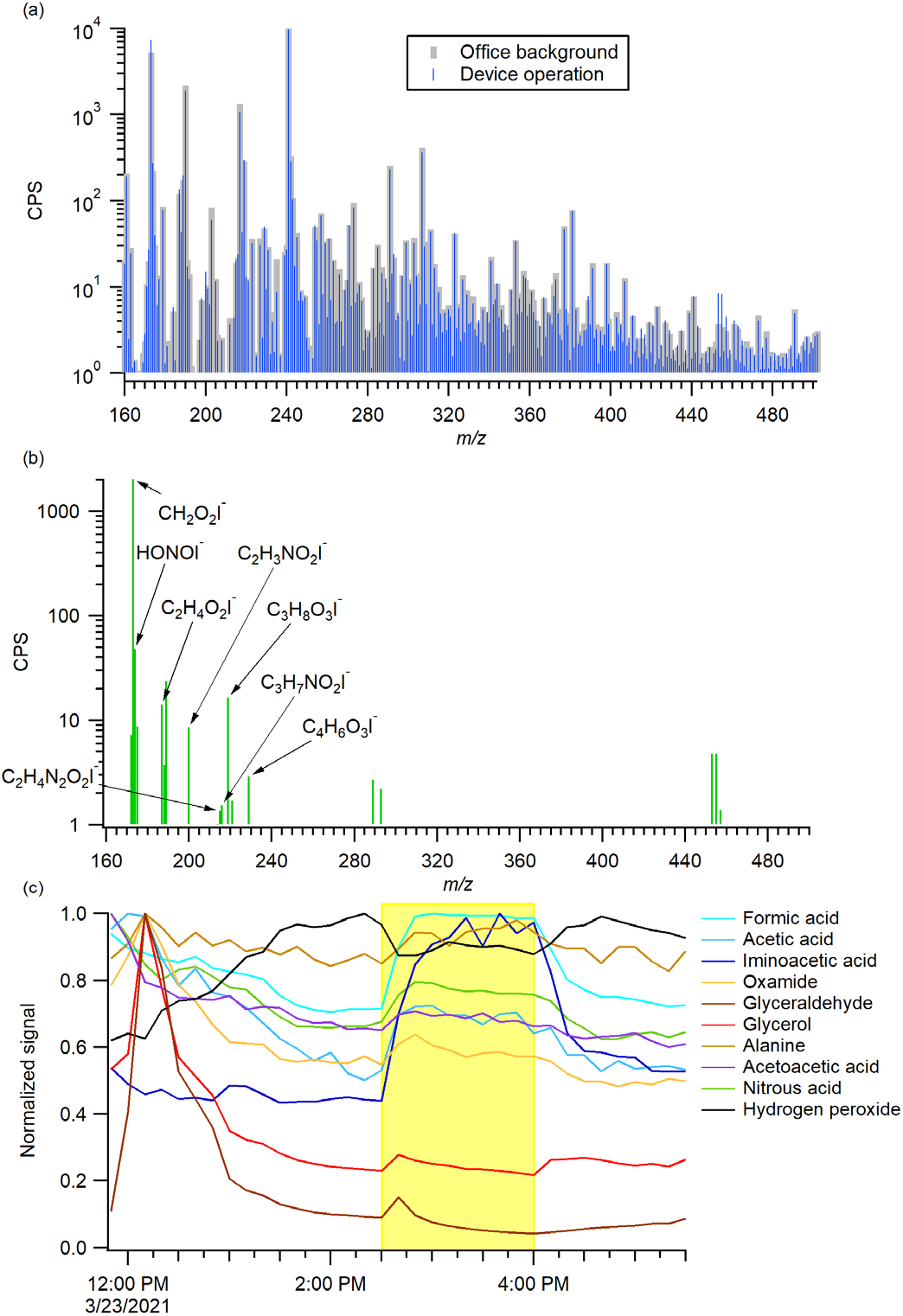
HR-ToF-CIMS results showing (a) grey: office background mass spectrum / blue: mass spectrum during hydroxyl generator operation, (b) the mass spectrum difference between before and during the operation of hydroxyl generator, and (c) time evolution of selected species. The data are 10-min averaged data and are normalized by the maximum signal of each species. The hydroxyl generator was in operation from 2:30 pm to 4:00 pm (highlighted in yellow). Glyceraldehyde (*m/z* 217, C_3_H_6_O_3_I^-^) is not included in the mass spectrum in (b) due to its rapid decay after the formation.

### Formation of secondary organic aerosol

Particle number and volume concentrations started increasing once the device was in operation (Figure 2a). Both the number and volume concentrations increased rapidly in the first 30 minutes after the device was turned on and slowed down after reaching ∼4000 particle cm^-3^ and ∼5 µm^3^ cm^-3^, respectively. This was followed by a rapid decrease in concentrations after the device was turned off. The increase in particles was mostly within the PM_1_ size range based on the agreement between the SMPS (PM_1_ only) and the CPC (all particles under roughly 3 µm). During the operation, an enhancement was observed in the 100 - 200 nm size range for both particle number and volume concentrations (Figure S1). It is noted that a similar experiment was performed in a laboratory space (∼ 140 m^2^) and similar results were observed (Figure S2).

**Figure 2.**
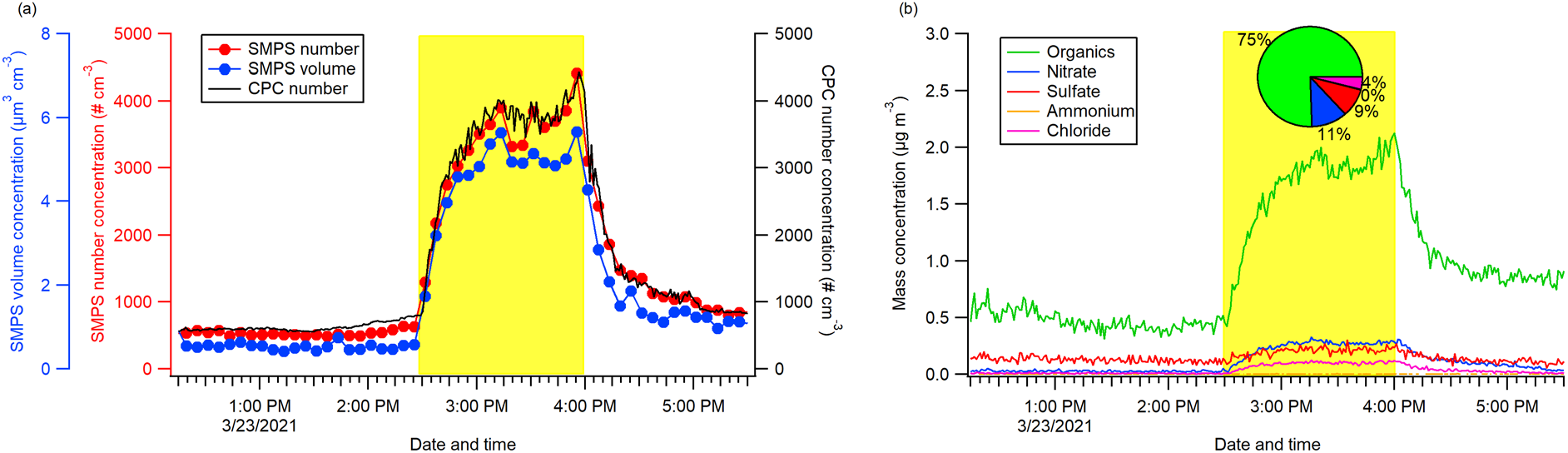
Time series of (a) particle number (CPC and SMPS) and volume concentrations (SMPS) and (b) non-refractory species concentrations (HR-ToF-AMS). The mass fraction of different non-refractory species during the hydroxyl generator operation is shown in the pie chart. The hydroxyl generator was in operation from 2:30 pm to 4:00 pm (highlighted in yellow).

The time series of the species measured by the HR-ToF-AMS are shown in Figure 2b. A collection efficiency of 0.45 was applied to the data as the inorganic concentrations were low and the aerosol did not contain high mass fractions of acidic sulfate or ammonium nitrate.^39^ The chemical composition of the particles during the operation period confirmed SOA formation, with organics reaching 2.1 μg m^-3^ after the device was turned on. Figure 2 also shows that the mass concentrations of non-refractory species reported by the HR-ToF-AMS were somewhat lower than the volume concentration enhancement measured by the SMPS (converted to mass concentration by the density of each species, Figure S3). This difference was expected since both instruments sampled particles without the use of a dryer at the instrument inlets. Thus, the particle concentration reported by the SMPS included water whereas particle water can be evaporated in the low-pressure aerodynamic lens and vacuum system of HR-ToF-AMS.^39-42^ The discrepancy diminishes when accounting for particle water as shown in Figure S3. Ammonium concentration was low throughout the experiment and showed little changes whereas nitrate, sulfate, and chloride increased during the device operation period. Since we did not observe ammonium increasing along with nitrate, sulfate, or chloride, these species are likely in the form of organic nitrate, organic sulfate, and organic chloride. Organic mass spectra comparison shows enhanced fraction at *m/z* 44 (CO_2_^+^) during the device operation (Figures 3a and S4), with increased O:C and decreased H:C (Figure S5a). The increase in the degree of oxidation of aerosol is further illustrated in the Van Krevelen diagram in Figure 3b.^43-45^ The aerosol evolution followed a slope of ∼ -1, with particle carbon oxidation state (OS_C_ = 2 O:C - H:C) increasing during device operation, as a result of enhancements in O:C and reductions in H:C.^46^

**Figure 3.**
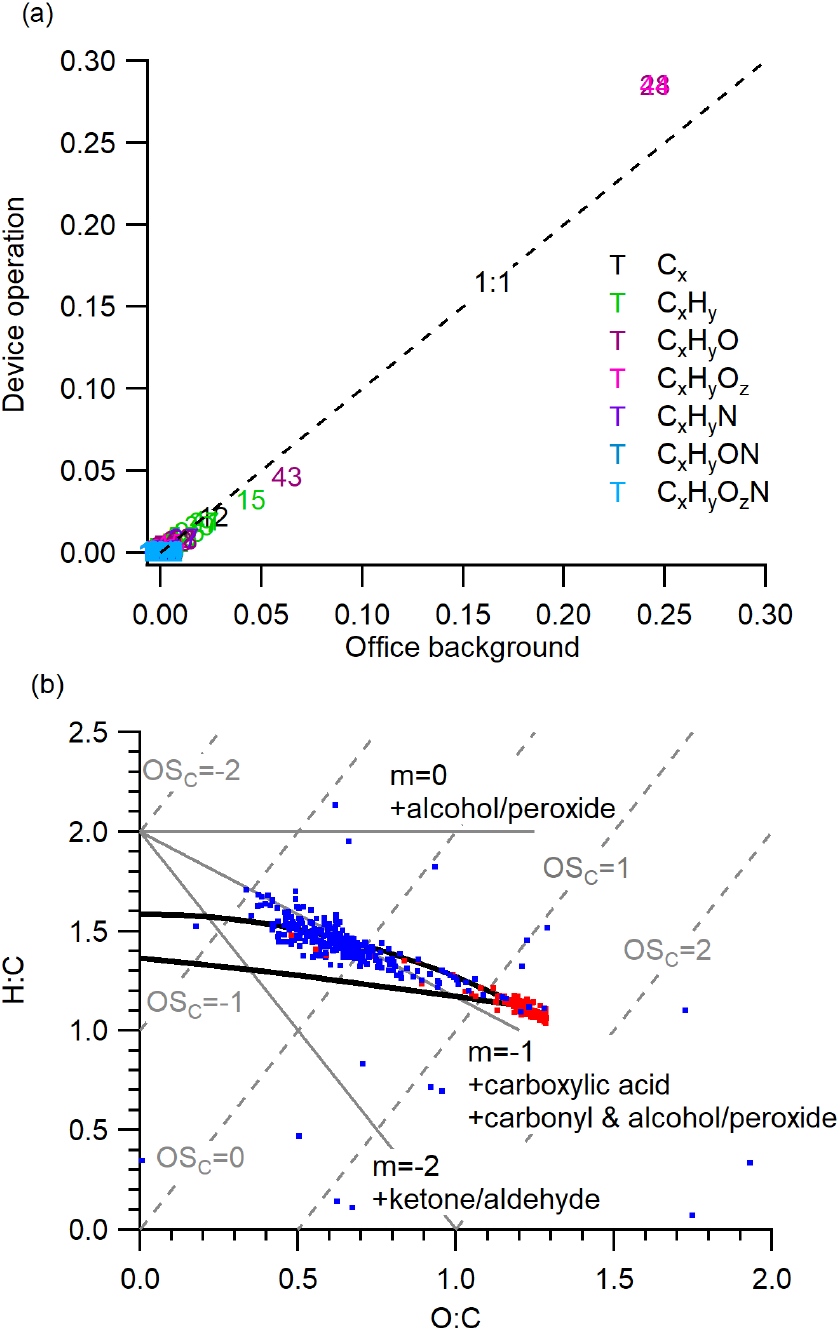
HR-ToF-AMS results showing (a) organic mass spectra comparison between during device operation and office background and (b) VK-triangle diagram of organics. The black lines encompass the triangular space occupied by ambient SOA.^44^ The carbon oxidation states (OS_C_) are shown with grey dotted lines. The blue data points correspond to office background and the red data points correspond to device operation.

## Discussion

Generation of hydroxyl radicals indoors reduces VOC concentrations in a similar manner to tropospheric VOC oxidation chemistry, which proceeds through complex, multi-generational chemistry and results in the formation of a large number of organic products. The byproducts formed from these reactions in this study depend on the identity of the VOCs in the office. VOCs were not measured in this work, however, Price et al.^47^ speciated total organic carbon (in both gas and particle phases) in an art museum and identified that over 80% of the carbon present are highly reduced (OS_C_ < -0.5) and volatile compounds with low carbon number (< C_6_). It is possible that the office has a similar VOC speciation, though it could have a lower total carbon budget due to no occupancy and no activities, such as printing,^48^ which can lead to VOC emissions. The observed oxygenated C_1_-C_4_ compounds can be formed from functionalization and fragmentation of such VOCs during the oxidation.^22^ Although we only observed small carboxylic acids in the gas phase, this does not exclude the formation of larger OVOCs, which might not be detected by I-CIMS, be lost in the instrument inlet line, interact with the surfaces,^1, 47^ or have participated in new particle formation and growth.^49^ Increasing particle number and mass concentrations and the formation of highly-oxidized SOA suggest that new particle formation and condensation growth can be a loss process of larger, less-volatile OVOCs. The SOA formed has an O:C of ∼ 1.3, which is higher than the typical O:C range observed for more-oxidized oxygenated organic aerosol (MO-OOA) in ambient environments.^44, 50^ While both nucleation and gas-particle partitioning can lead to SOA formation, nucleation is likely the main process as particle elemental composition changed (O:C decreased and H:C increased) as soon as the device was turned off. The prevalence of nucleation is likely due to the small condensation sink with low aerosol background (∼581 # cm^-3^) in the office. The enhancement of *m/z* 44 (CO_2_^+^) in the HR-ToF-AMS organic mass spectra indicates the contribution of organic acids in SOA formation, as their thermal decarboxylation gives rise to the CO_2_^+^ fragment.^44, 51-54^ In the Van Krevelen diagram, the SOA evolved along the ∼ -1 line, which corresponds to the addition of carboxylic acids and/or simultaneous increases in alcohol and carbonyl groups. ^43, 44^ Taken together, these results show that carboxylic acids were formed during the oxidation process and contributed to new particle formation owing to their low volatility. ^22, 49,55^

Nitrate, sulfate, and chloride enhancements are expected to be associated with organic nitrate, organic sulfate, and organic chloride formation (Figure S5). The average NO^+^/NO_2_^+^ ion ratio from the HR-ToF-AMS is widely used as an indicator to differentiate inorganic vs. organic nitrate.^56-58^ The NO^+^/NO_2_^+^ ratio for inorganic nitrate during the instrument calibration was 1.98, and previous laboratory studies have shown that this ratio is much higher for organic nitrate than inorganic nitrate.^56, 59-61^ The average NO^+^/NO_2_^+^ ratio during the operation period was ∼17, implying that virtually all the particle-phase nitrate was organic nitrate. The contribution of organic sulfate can be examined by evaluating the fractions of HSO_3_^+^ and H_2_SO4^+^ in H_x_SO_y_^+^ fragments (SO^+^, SO_2_^+^, SO_3_^+^, HSO_3_^+^, and H_2_SO4^+^).^62^ Both fractions decreased when the device was turned on, implying the presence of organic sulfate. Organic chloride formation can be associated with chlorine-containing VOCs which may be emitted or formed through interactions with cleaning products.^63,64^

To our knowledge, this is the first study that monitored the chemical composition of secondary products in both gas and particle phases during the operation of an electronic air cleaner that dissipates oxidants in a real-world setting. Although we lack parent VOC measurements, the limited number of OVOCs and small enhancement in SOA observed during this work were assumed to be due to low initial VOCs concentrations in the office where this study was conducted. Much larger enhancements in OVOCs and SOA could be observed in other types of indoor environments such as industrial settings, homes, and restaurants, which can have much larger VOC concentrations, even more than in outdoor locations. ^1, 47, 64-66^ Secondary VOC oxidation products have been shown to have detrimental effects on human health. ^24, 26, 27, 67, 68^ Specifically, SOA has been reported to induce cellular reactive oxygen species (ROS) generation, inflammatory cytokine production, and oxidative modification of RNA.^69-71^ The toxicity of SOA could increase with increasing OS_C_.^28, 72^ Therefore, future studies on air cleaning technologies should not be limited to the inactivation of bioaerosol or reduction of particular VOCs, but should also evaluate potential OVOCs and SOA formation during their operation. The electronic air cleaner tested in this study is similar to many other commercially available devices and similar experiments should be conducted with other devices.

## Data Availability

All data is available in the main text or the supporting information.

## Acknowledgements

The authors would like to thank C. Peng, A. P. Mouat, and J. Kaiser for helpful discussions on the experimental setup and J. Lee for help in the table of content (TOC) figure illustration. The HR-ToF-CIMS was purchased through NSF Major Research Instrumentation (MRI) grant 1428738.

## For TOC only

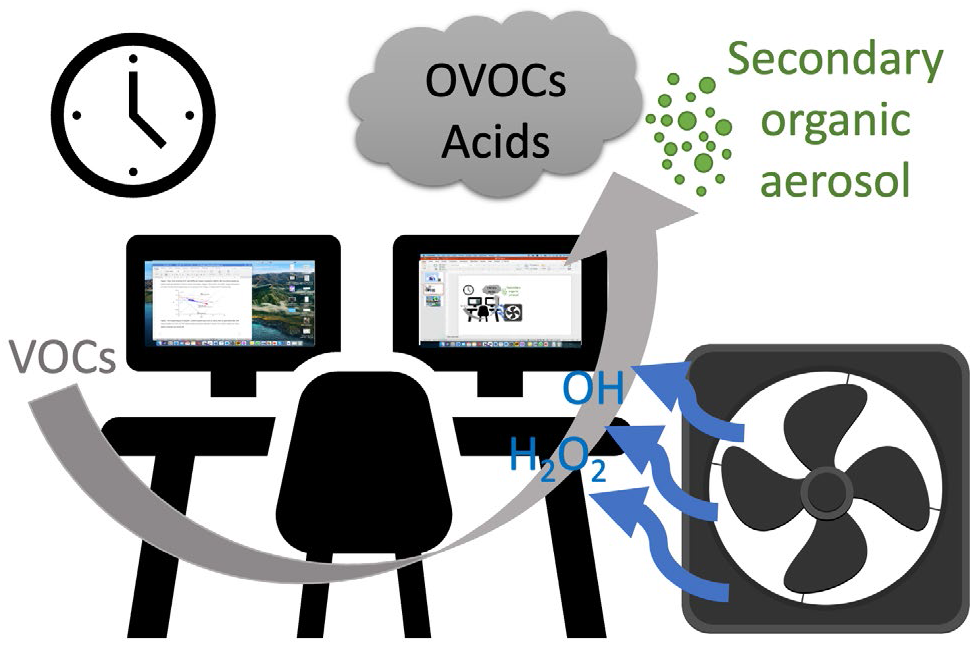

