## Supporting information for "Formation of oxidized gases and secondary organic aerosol from a commercial oxidant-generating electronic air cleaner"

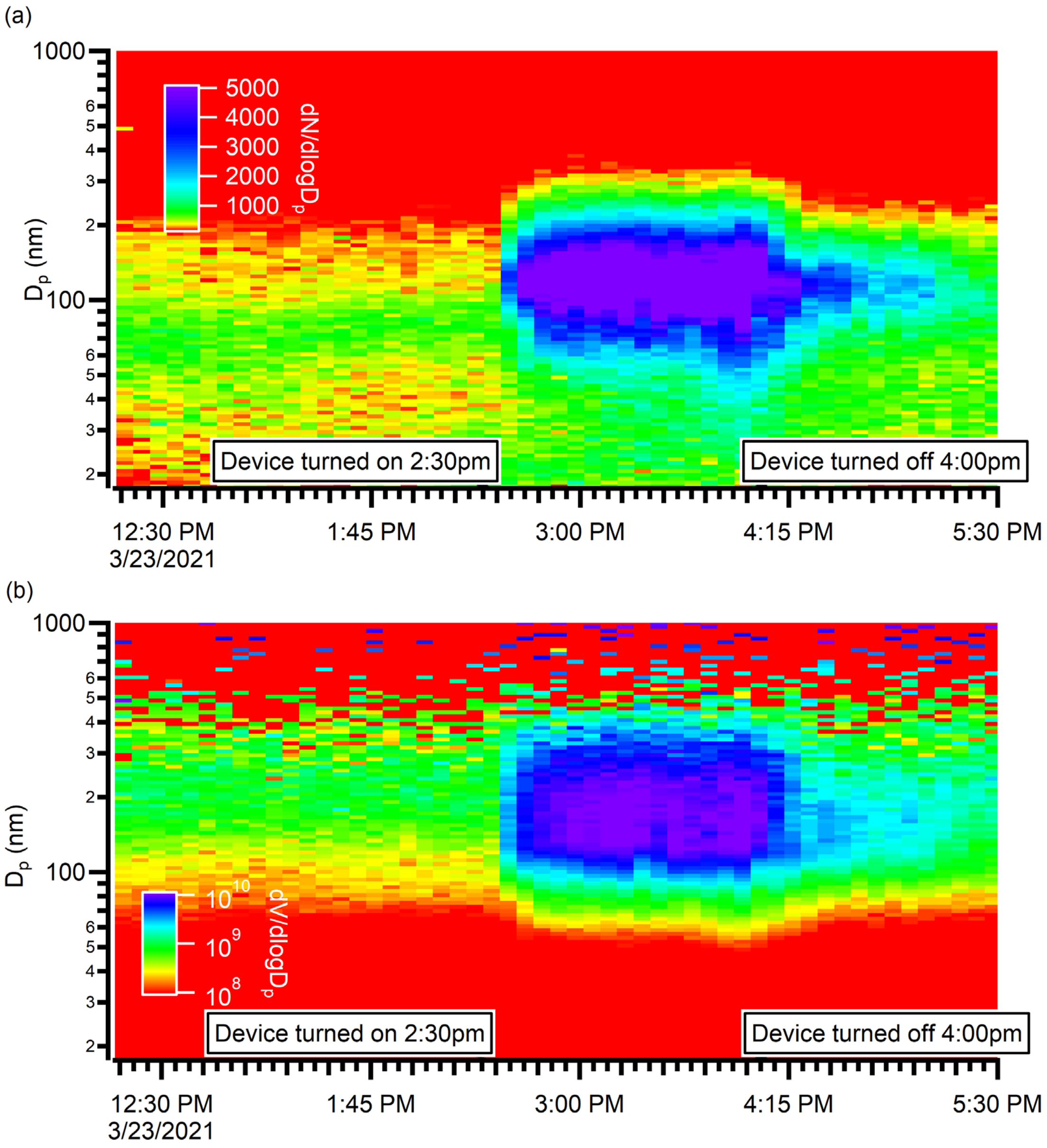


Figure S1. Size distribution of (a) particle number concentration (# $cm^{-3}$) and (b) particle volume concentration ($nm^3$ $cm^{-3}$) from the office measurements. The color scale is in log scale.

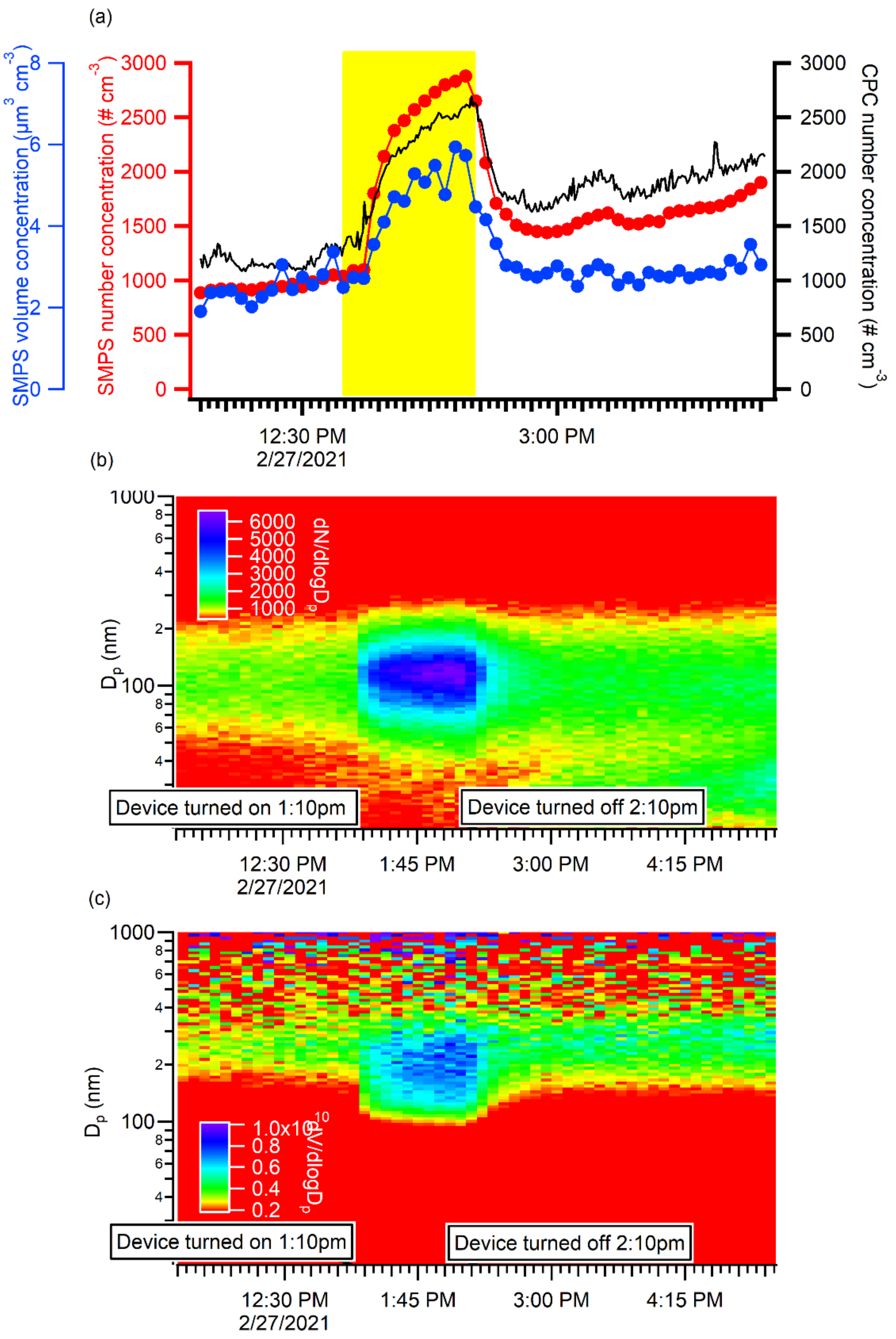


Figure S2. SMPS and CPC results from an experiment performed in a larger space (laboratory) than the office: (a) time series of number (CPC and SMPS) and volume concentrations (SMPS) with device operation period highlighted in yellow. Size distribution of (b) particle number concentration (# $cm^{-3}$) and (c) particle volume concentration ($nm^3$ $cm^{-3}$). The color scale is in log scale.

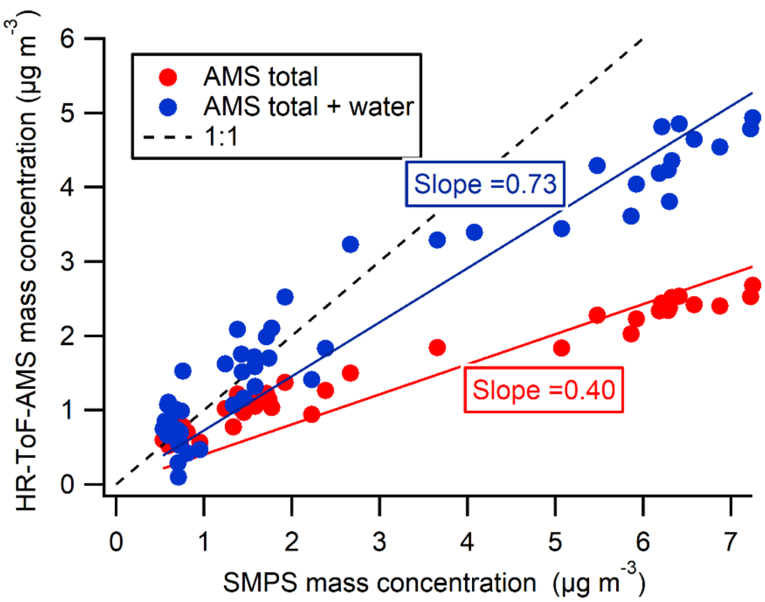


Figure S3. Comparison of AMS total mass concentration (sum of all non-refractory species) and SMPS mass concentration (intercepts set to zero) from the office measurements. The SMPS volume concentration was converted into mass concentration using density of the individual species: 1.75 g/cm$^3$ for sulfate, nitrate, and ammonium; 1.52 g/cm$^3$ for chloride; 1.79 g cm$^{-3}$ for organics, estimated based on the elemental ratio of organics.[1] Particle water from AMS was estimated based on the equation proposed in Engelhart et al.[2]

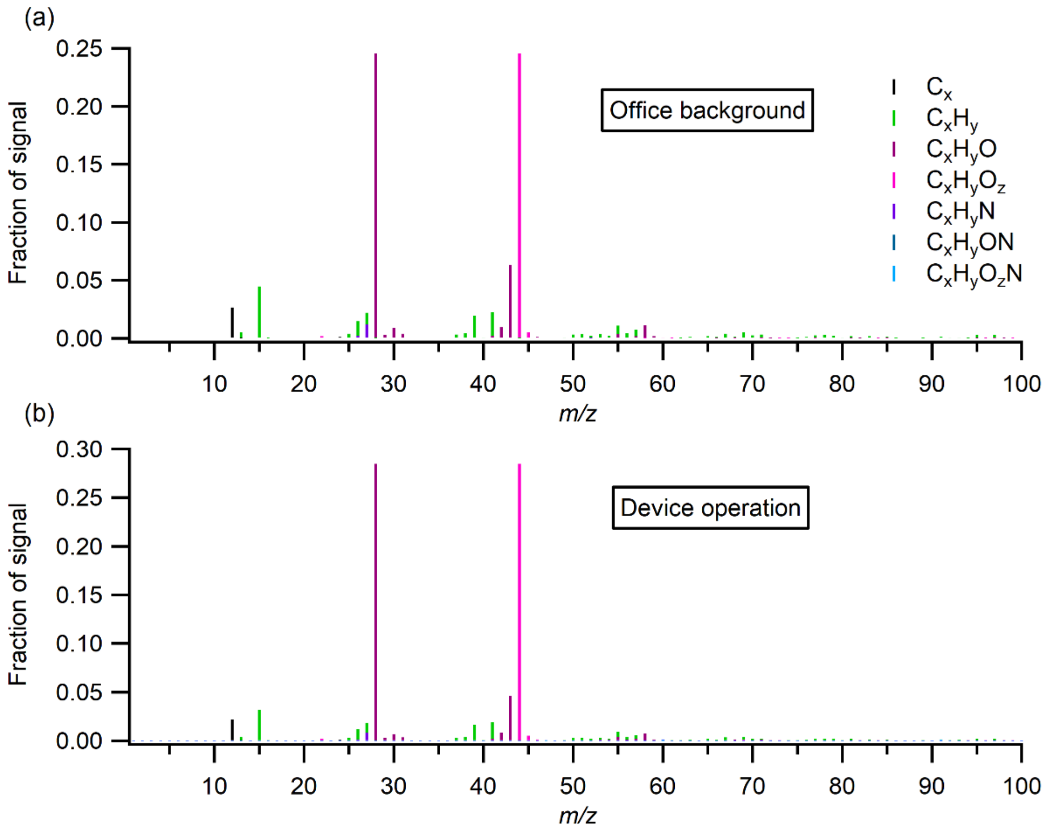


Figure S4. Organic families mass spectra of (a) office background and (b) during device operation.

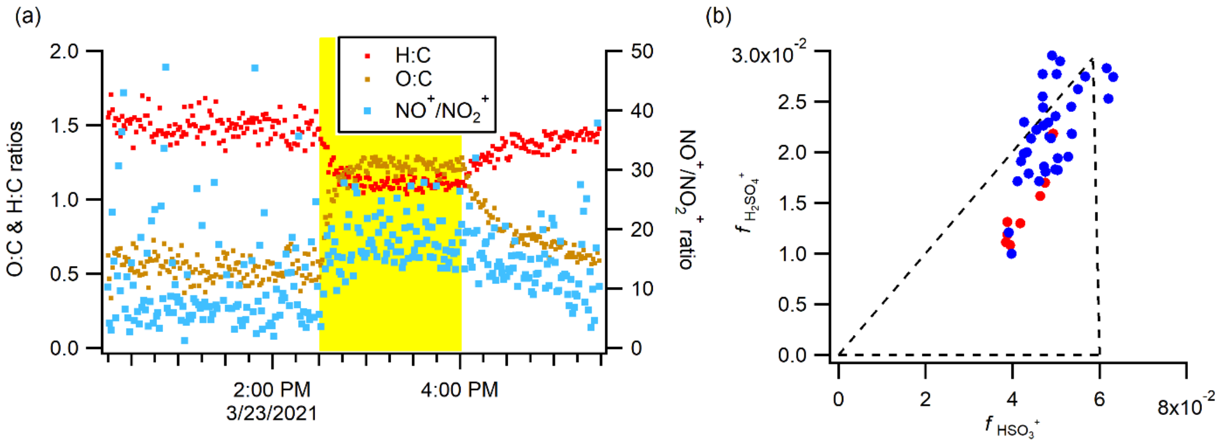


Figure S5. (a) Time series of H:C, O:C, and $NO^+/NO_2^+$ ratio from the office measurements, with the device operation period highlighted in yellow. (b) $f_{H2SO4+}$ vs. $f_{HSO3+}$ during the experiment. $f_{H2SO4+}$ and $f_{HSO3+}$ are the fractions of $H_2SO_4^+$ and $HSO_3^+$ in $H_xSO_y^+$ fragments ($SO^+$, $SO_2^+$, $SO_3^+$, $HSO_3^+$, and $H_2SO4^+$), respectively. Both fractions approach zero when organic sulfate contribution to total sulfate increases.[3] Data are 10-min averaged data. The blue data points correspond to office background and the red data points correspond to device operation. The two blue data points at around [0.04, 0.01] are the data points immediately after the device was turned off.

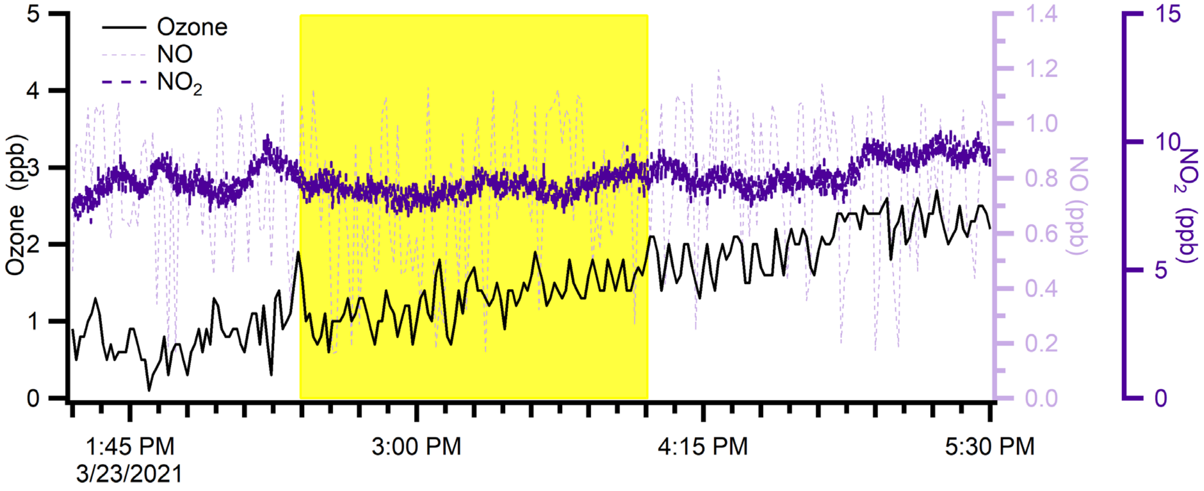


Figure S6. Time series of ozone, NO, and $NO_2$ from the office measurements. The device was in operation from 2:30 pm to 4:00 pm (highlighted in yellow).
